# Parents’ Likelihood to Vaccinate Their Children and Themselves Against COVID-19

**DOI:** 10.1101/2020.11.10.20228759

**Authors:** Matthew M. Davis, Joseph S. Zickafoose, Alese E. Halvorson, Stephen W. Patrick

## Abstract

**Background:** Vaccination against COVID-19 will likely involve children in order to mitigate transmission risks in community settings. Successful implementation of COVID-19 immunization in the United States may hinge on factors associated with parents’ likelihood of immunizing their children and themselves.

**Methods:** We fielded a national household survey in English and Spanish from June 5-10, 2020 (n=1,008). Parents were asked about their likelihood of immunizing their children and themselves against COVID-19. We fit separate regression models of parents’ likelihood to vaccinate themselves and their children against COVID-19, using bivariate and multivariable approaches in analyses weighted to be nationally representative.

**Results:** Overall, 63% of parents (95% CI: 59%, 66%) were likely to vaccinate their children against COVID-19, and 60% (57%, 64%) were likely to get a vaccine themselves. These responses were highly correlated (Pearson’s r=0.89). Parent age, sex, marital status, education level, and income were all associated with parents’ likelihood to vaccinate their children and themselves in bivariate analyses; race/ethnicity was significantly associated with parents’ likelihood to vaccinate their children. In multivariable analyses, younger parents were significantly less likely than older parents to vaccinate their children and themselves against COVID-19, as were parents with high school or less education compared with parents with bachelor’s degrees and non-Hispanic White parents compared with Hispanic parents (all p<.05).

**Conclusion:** In this national survey, only approximately 60% of U.S. parents stated that they are likely to vaccinate their children or themselves against COVID-19. Addressing parents’ hesitancy to vaccinate themselves and their children against COVID-19 will be instrumental to achieving herd immunity in the US.

## Introduction

Vaccination against COVID-19 will likely involve children in order to mitigate transmission risks in school and daycare settings.^1^ Successful implementation of a COVID-19 immunization program in the United States will depend on factors commonly associated with vaccination patterns, such as sociodemographic characteristics,^2^ and may also relate to environmental exposures such as COVID-19 case incidence and mortality rates. No peer-reviewed studies have examined parents’ likelihood to vaccinate their children and themselves against COVID-19.

## Methods

To assess factors associated with parents’ likelihood to vaccinate themselves and their children against COVID-19, we fielded a national household survey in English and Spanish from June 5-10, 2020.^3^ The survey had a 50% completion rate (n=1,008);^4^ survey weights were assigned to permit national estimates of the attitudes of parents with at least 1 child younger than age 18 while accounting for differential non-response. This study was considered exempt from human subjects review by the Vanderbilt University Medical Center Institutional Review Board.

Parents were asked two vaccination-specific questions (“If a vaccine against COVID-19 becomes available in the next 12 months, how likely are you to get it for [yourself/your child(ren)]?”). Four original response options were dichotomized into “Likely” (“Very likely” and “Somewhat likely”) and “Unlikely” (“Not too likely” and “Not at all likely”) for purposes of analysis. We abstracted state-level COVID-19 cumulative case counts and case fatality rates (https://coronavirus.jhu.edu/data/state-timeline) in the middle of our survey field period (June 8) as well as changes from 1 month prior (May 8). We fit separate models of parents’ likelihood to vaccinate themselves and their children against COVID-19, using bivariate and multivariable approaches in R version 3.6.2 (R Core Team, Vienna, Austria).

## Results

Overall, 63% of parents (95% CI: 59%, 66%) were likely to vaccinate their children against COVID-19, and 60% (57%, 64%) were likely to get a vaccine themselves. These responses were highly correlated (Pearson’s r=0.89). Parent age, sex, marital status, education level, and income were all associated with parents’ likelihood to vaccinate their children and themselves in bivariate analyses; race/ethnicity was significantly associated with parents’ likelihood to vaccinate their children (**Table 1**). Parent employment status, child age, and state-level population-adjusted COVID-19 case incidence and mortality rates were not associated with likelihood to vaccinate (data not shown).

**Table 1.** Parents’ likelihood of vaccinating their child(ren) and themselves against COVID-19, by sociodemographic characteristics.

| **Sociodemographic Variables**<br>(unweighted n) | Parents' likelihood of vaccinating their child(ren)* |  |  | Parents' likelihood of vaccinating themselves* |  |  |
| --- | --- | --- | --- | --- | --- | --- |
|  | Likely<sup>a</sup> | Unlikely<sup>a</sup> | p-value** | Likely<sup>a</sup> | Unlikely<sup>a</sup> | p-value** |
| *Overall (n=1,008)* | 62.8% (59.5, 66.1) | 37.2% (33.9, 40.5) |  | 60.4% (57.1, 63.7) | 39.6% (36.3, 42.9) |  |
| **Parent Age** |  |  | p<0.001 |  |  | p<0.001 |
| 18-35 (n=230) | 52.1% (45.2, 58.9) | 47.9% (41.1, 54.8) |  | 49.9% (43.0, 56.8) | 50.1% (43.2, 57.0) |  |
| 36-45 (n=460) | 67.1% (62.6, 71.6) | 32.9% (28.4, 37.4) |  | 64.8% (60.2, 69.4) | 35.2% (30.6, 39.8) |  |
| 46+ (n=318) | 69.3% (64.0, 74.6) | 30.7% (25.4, 36.0) |  | 66.6% (61.1, 72.1) | 33.4% (27.9, 38.9) |  |
| **Parent Sex** |  |  | p=0.003 |  |  | p=0.039 |
| Male (n=453) | 68.3% (63.6, 73.0) | 31.7% (27.0, 36.4) |  | 64.3% (59.5, 69.2) | 35.7% (30.8, 40.5) |  |
| Female (n=555) | 58.4% (53.8, 62.9) | 41.6% (37.1, 46.2) |  | 57.3% (52.8, 61.8) | 42.7% (38.2, 47.2) |  |
| **Parent Marital Status** |  |  | p=0.001 |  |  | p<0.001 |
| Married (n=838) | 65.6% (62.1, 69.1) | 34.4% (30.9, 37.9) |  | 64.3% (60.7, 67.8) | 35.7% (32.2, 39.3) |  |
| Unmarried (n=170) | 51.1% (42.8, 59.4) | 48.9% (40.6, 57.2) |  | 44.3% (36.1, 52.6) | 55.7% (47.4, 63.9) |  |
| **Parent Education** |  |  | p<0.001 |  |  | p<0.001 |
| High school or less (n=312) | 55.1% (49.1, 61.1) | 44.9% (38.9, 50.9) |  | 52.6% (46.6, 58.6) | 47.4% (41.4, 53.4) |  |
| Some college (n=230) | 55.0% (48.0, 61.9) | 45.0% (38.1, 52.0) |  | 51.8% (44.8, 58.8) | 48.2% (41.2, 55.2) |  |
| College degree or more (n=466) | 74.2% (69.9, 78.6) | 25.8% (21.4, 30.1) |  | 72.4% (68.0, 76.9) | 27.6% (23.1, 32.0) |  |
| **Parent Race/Ethnicity** |  |  | p=0.036 |  |  | p=0.087 |
| White, Non-Hispanic (n=661) | 60.9% (56.9, 64.9) | 39.1% (35.1, 43.1) |  | 59.2% (55.2, 63.2) | 40.8% (36.8, 44.8) |  |
| Black, Non-Hispanic (n=78) | 54.7% (43.0, 66.4) | 45.3% (33.6, 57.0) |  | 51.3% (39.6, 63.1) | 48.7% (36.9, 60.4) |  |
| Other, Non-Hispanic (n=91) | 76.3% (65.9, 86.7) | 23.7% (13.3, 34.1) |  | 71.0% (60.0, 82.0) | 29.0% (18.0, 40.0) |  |
| Hispanic (n=178) | 65.4% (57.7, 73.1) | 34.6% (26.9, 42.3) |  | 63.1% (55.3, 70.8) | 36.9% (29.2, 44.7) |  |
| **Household Income** |  |  | p<0.001 |  |  | p<0.001 |
| Less than \$25,000 (n=86) | 47.9% (36.5, 59.4) | 52.1% (40.6, 63.5) | | 42.9% (31.6, 54.2) | 57.1% (45.8, 68.4) | |
| \$25,000 to \$49,999 (n=138) | 48.4% (39.3, 57.4) | 51.6% (42.6, 60.7) | | 45.3% (36.2, 54.4) | 54.7% (45.6, 63.8) | |
| \$50,000 to \$99,999 (n=313) | 64.5% (58.6, 70.3) | 35.5% (29.7, 41.4) | | 62.5% (56.6, 68.3) | 37.5% (31.7, 43.4) | |
| \$100,000 or more (n=471) | 69.9% (65.4, 74.5) | 30.1% (25.5, 34.6) | | 68.1% (63.5, 72.7) | 31.9% (27.3, 36.5) | |

<sup>a</sup> Based on parents' response to the questions: "If a vaccine against COVID-19 becomes available in the next 12 months, how likely are you to get it for [yourself/your child(ren)]?". Response options were: Very likely / Somewhat likely / Not too likely / Not at all likely. For analysis, we collapsed "Very likely" and "Somewhat likely" as "Likely" and "Not too likely" and "Not at all likely" as "Unlikely".
\* Reported as weighted proportions with 95% confidence intervals.
\*\* p-values reported from Rao-Scott corrected chi-square tests

In multivariable analyses, older parents were significantly more likely than younger parents to vaccinate their children and themselves against COVID-19, as were parents with bachelor’s degrees versus parents with high school or less education and Hispanic parents compared with non-Hispanic White peers (**Figure 1**).

**Figure 1.**
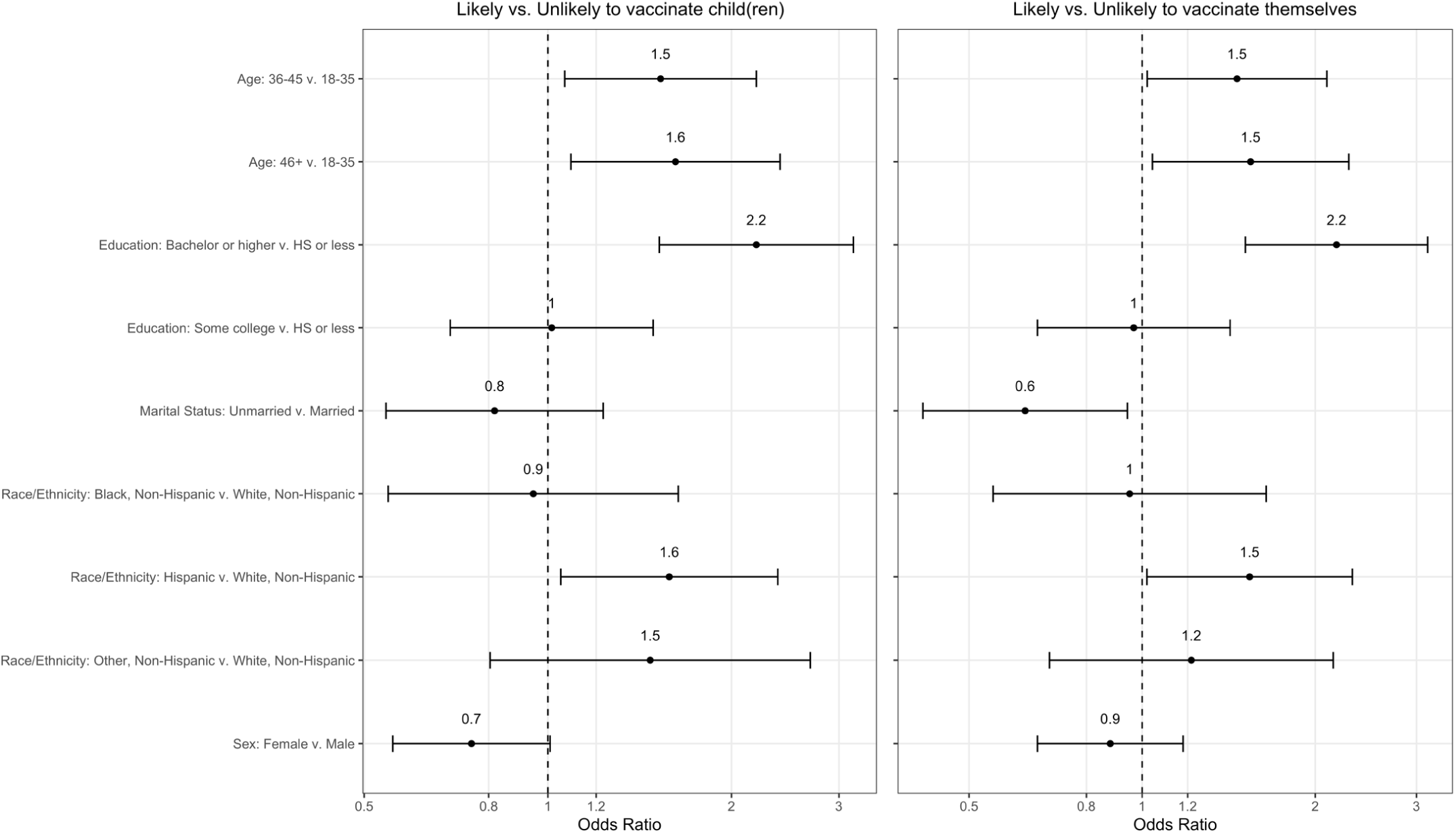
Multivariable models of association of sociodemographic factors with parents’ likelihood of vaccinating their child(ren) and themselves against COVID-19. Household income was not included in the model because of collinearity with parent education.

## Discussion

In this national survey, only approximately 60% of U.S. parents stated that they are likely to vaccinate their children or themselves against COVID-19. This level of hesitancy among US adults about vaccination against COVID-19 is similar to a separate study among adults (not limited to parents with minor children) fielded about 6 weeks earlier, which also found associations of hesitancy with younger adult age and lower educational attainment.^2^

Greater hesitancy regarding childhood vaccines is also commonly associated with less education among parents, often related to strong belief systems regarding risks of disease versus risks of inoculation.^5,6^ Our study may be limited by a sample that included a disproportionately high number of high-income respondents, although this is similar to other recent national surveys of this topic.^5^ We attempted to mitigate this sampling bias through population-based survey weighting in our analyses. Addressing parents’ hesitancy to vaccinate themselves and their children against COVID-19 will likely be essential to achieve herd immunity. Working with parents to understand personal reasons for hesitancy will help facilitate uptake when safe and effective vaccines to prevent COVID-19 become available.

## Data Availability

These data are not publicly available.

## Acknowledgements

The authors would like to thank Seethalakshmi Davis for her contributions to data collection for this project.

## Conflict of Interest Disclosures

The authors have no conflicts of interests to disclose.

## Funding/Support

None.

## Role of the Sponsor

Not applicable.

## Notes

### Competing Interest Statement

The authors have declared no competing interest.

### Author Declarations

This study was considered exempt from human subjects review by the Vanderbilt University Medical Center Institutional Review Board.

